# Systematic review of genotype-stratified treatment for monogenic insulin resistance

**DOI:** 10.1101/2023.04.17.23288671

**Authors:** Robert K. Semple, Kashyap A. Pate, Sungyoung Auh, ADA/EASD PMDI, Rebecca J. Brown

## Abstract

**Objective:** To assess the effects of pharmacologic and/or surgical interventions in monogenic insulin resistance (IR), stratified by genetic aetiology.

**Design:** Systematic review.

**Data sources:** PubMed, MEDLINE and Embase, from 1 January 1987 to 23 June 2021.

**Review methods:** Studies reporting individual-level effects of pharmacologic and/or surgical interventions in monogenic IR were eligible. Individual subject data were extracted and duplicate data removed. Outcomes were analyzed for each affected gene and intervention, and in aggregate for partial, generalised and all lipodystrophy.

**Results:** 10 non-randomised experimental studies, 8 case series, and 21 single case reports met inclusion criteria, all rated as having moderate or serious risk of bias. Metreleptin was associated with lower triglycerides and hemoglobin A1c in aggregated lipodystrophy (n=111), in partial lipodystrophy (n=71) and generalised lipodystrophy (n=41)), and in *LMNA*, *PPARG*, *AGPAT2* or *BSCL2* subgroups (n=72,13,21 and 21 respectively). Body Mass Index (BMI) was lower after treatment in partial and generalised lipodystrophy overall, and in *LMNA or BSCL2*, but not *PPARG* or *AGPAT2* subgroups. Thiazolidinedione use was associated with improved hemoglobin A1c and triglycerides in aggregated lipodystrophy (n=13), improved hemoglobin A1c only in the *PPARG* subgroup (n=5), and improved triglycerides only in the *LMNA* subgroup (n=7). In *INSR*-related IR, use of rhIGF-1, alone or with IGFBP3, was associated with improved hemoglobin A1c (n=15). The small size or absence of all other genotype-treatment combinations precluded firm conclusions.

**Conclusions:** The evidence guiding genotype-specific treatment of monogenic IR is of low to very low quality. Metreleptin and Thiazolidinediones appear to have beneficial metabolic effects in lipodystrophy, and rhIGF-1 appears to lower hemoglobin A1c in INSR-related IR. For other interventions there is insufficient evidence to assess efficacy and risks either in aggregated lipodystrophy or in genetic subgroups. There is a pressing need to improve the evidence base for management of monogenic IR.

## Introduction

Diabetes caused by single gene changes is highly heterogeneous in molecular aetiopathogenesis. It may be grouped into disorders featuring primary failure of insulin secretion, and disorders in which insulin resistance (IR), often severe, predates secondary failure of insulin secretion and diabetes. Monogenic IR is itself heterogeneous, encompassing primary lipodystrophy syndromes, primary disorders of insulin signalling, and a group of conditions in which severe IR is part of a more complex developmental syndrome 1.

Monogenic IR is rare but underdiagnosed. The commonest subgroup is formed by genetic lipodystrophy syndromes ^2, 3^. Recent analysis of a large clinical care cohort unselected for metabolic disease suggested a clinical prevalence of lipodystrophy of around 1 in 20,000, with a prevalence of plausible lipodystrophy-causing genetic variants of around 1 in 7,000 4. Monogenic IR is important to recognise, because affected patients are at risk not only of micro- and macrovascular complications of diabetes, but also of complications such as dyslipidemia, pancreatitis, and steatohepatitis, especially in lipodystrophy syndromes 5. Non-metabolic complications specific to individual gene defects may also occur, including hypertrophic cardiomyopathy and other manifestations of soft tissue overgrowth 3. Diabetes is also commonly the sentinel presentation of a multisystem disorder, and recognition of complex syndromes in a diabetes clinic may trigger definitive diagnostic testing.

The only therapy licensed specifically for monogenic IR is recombinant human methionyl leptin (metreleptin), with licensed indications encompassing a subset of patients with lipodystrophy and inadequate metabolic control ^6, 7^. The current license in the USA is restricted to generalised lipodystrophy, but in Europe it extends to some patients with partial lipodystrophy. A significant proportion of the body of evidence considered in licensing addressed patients ascertained by presence of clinical lipodystrophy, and the role of genetic stratification in precision treatment of lipodystrophy has not been systematically addressed. Many other medications and other treatment options are also widely used in monogenic IR, although not licensed for that specific subgroup. Such use draws on the evidence base and treatment algorithms developed for type 2 diabetes. Several forms of monogenic IR have molecular and/or clinical attributes that suggest potential precision approaches to treatment.

We sought now to undertake a systematic review of the current evidence guiding treatment of monogenic IR stratified by genetic aetiology, to assess evidence for differential responses to currently used therapies, to establish gaps in evidence, and to inform future studies. This systematic review is written on behalf of the American Diabetes Association (ADA)/European Association for the Study of Diabetes (EASD) *Precision Medicine in Diabetes Initiative* (PMDI) as part of a comprehensive evidence evaluation in support of the 2^nd^ International Consensus Report on Precision Diabetes Medicine [Tobias et al, Nat Med]. The PMDI was established in 2018 by the ADA in partnership with the EASD to address the burgeoning need for better diabetes prevention and care through precision medicine 8.

## Methods

### Inclusion Criteria and Search Methodology

To assess treatment of severe IR of known monogenic aetiology, with or without diabetes mellitus, including generalised and partial lipodystrophy and genetic disorders of the insulin receptor, we developed, registered and followed a protocol for a systematic review (Prospero ID CRD42021265365; registered July 21, 2021)9. The study was reported in accordance with Preferred Reporting Items for Systematic Reviews and Meta-Analysis (PRISMA) guidelines. Filtering and selection of studies for data extraction were recorded using the Covidence platform (https://www.covidence.org, Melbourne, Australia).

We searched PubMed, MEDLINE and Embase from 1987 (the year before identification of the first monogenic aetiology of IR) to June 23, 2021 for potentially relevant human studies in English. We used broad search terms designed to capture the heterogeneity of monogenic IR and its treatments. We searched for studies addressing 1. Severe IR due to variant(s) in a single gene OR 2. Congenital generalised or familial partial lipodystrophy due to variant(s) in a single gene. We selected only studies that reported a treatment term, including but not limited to mention of 1. Thiazolidinediones (TZD), 2. Metreleptin, 3. SGLT2 inhibitors, 4. GLP-1 analogues, 5. Bariatric surgery (all types), 6. Recombinant human IGF-1 or IGF-1/IGFBP3 composite, 7. U-500 insulin. No interventions were excluded in the primary search. In addition to the automated search, we hand searched reference lists of relevant review articles. Given the rarity of monogenic IR, no study types were excluded in the initial search. We ultimately considered experimental studies, case reports, and case series. The full search strategy is described in Supplemental Table 1.

Study selection for data extraction was performed in two phases, namely primary screening of title and abstract, then full text review of potentially eligible articles. Two authors independently evaluated eligibility, with discrepancies resolved by a third investigator. We excluded publications without original data, such as reviews, editorials, and comments, and those solely addressing severe IR or lipodystrophy of unknown or known non-monogenic aetiology, including HIV-related or other acquired lipodystrophies, or autoimmune insulin receptoropathy (Type B insulin resistance). Studies in which no clear categorical or numerical outcome of an intervention was reported, or in which interventions were administered for less than 28 days were also excluded.

### Data extraction and outcome assessments

One author extracted data from each eligible study using data extraction sheets. Data from each study was verified by all 3 authors to reach consensus. Data were extracted from text, tables, or figures. Study investigators were contacted for pertinent unreported data or additional details where possible, most commonly genetic aetiology of insulin resistance in reported patients, and outcome data.

Data extracted for each study included first author, publication year, country, details of intervention, duration of follow-up, study design, and number of participants. Subject-level data were extracted for outcomes of interest, including sex, genetic cause of severe insulin resistance (gene name, mono- vs biallelic *INSR* pathogenic variant), phenotypic details of severe IR/lipodystrophic subtype (generalised vs partial lipodystrophy; associated syndromic features). Subject level outcome data for were extracted prior to and after the longest-reported exposure to the intervention of interest for hemoglobin A1c (A1c), body mass index, serum triglyceride, ALT, or AST concentration, any index of liver size or lipid content, and total daily insulin dose. Potential adverse effects of interventions were recorded, including urinary tract infection, genital candidiasis, hypoglycemia, excessive weight loss, pancreatitis, soft tissue overgrowth, and tumor formation.

### Risk of bias and certainty of evidence assessment

Quality of extracted case reports and case series was assessed using NIH Study Quality Assessment Tools10. Grading of overall evidence for specific research questions was undertaken as detailed in 11.

### Data synthesis and analysis

Extracted data were managed using Covidence and analysed with SAS version 9.4. Pooled analysis was undertaken for all combinations of genotype and intervention for which sufficient numbers were reported, as well as for aggregated lipodystrophies, and generalized and partial subgroups of lipodystrophy. Generalized Estimating Equation models were used with time as a fixed factor and study as a random factor to examine treatment effects. Serum triglyceride concentrations were analyzed with and without log transformation. Data were summarized using estimated least-squared means with corresponding 95% confidence intervals.

## Results

### Identification of eligible studies

Initial searching identified 2,933 studies, to which 109 were added from the bibliography of a recent comprehensive review of monogenic lipodystrophy 2. 248 articles remained after screening of titles and abstracts, and 42 after full text screening (Figure 1).

**Figure 1:**
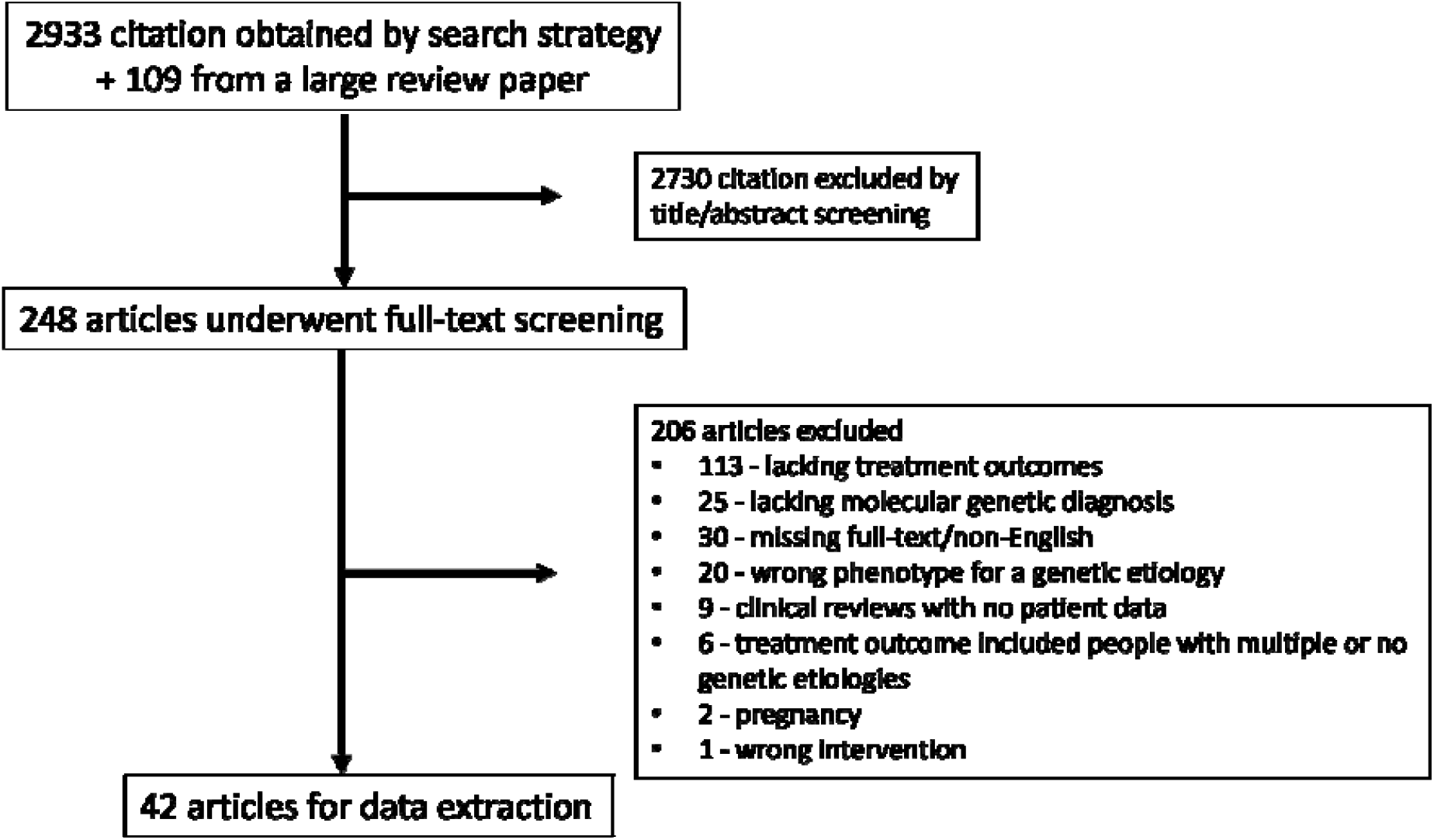
Flow diagram of publications evaluated based on the search strategy.

### Included studies addressed limited interventions and most had a high risk of bias

The 42 studies analysed, and assessment of their quality are summarised in Table 1 and detailed in Supplemental Table 2. Study quality was assessed as being fair in 15 cases and poor in 27 cases, including all case reports. This was primarily due to high risk of bias, particularly related to lack of control group for all studies. Three of the 42 studies included in further analysis included only individuals already described in other reports and were discarded, leaving 39 studies for final analysis. These comprised 10 non-controlled experimental studies, 8 case series and 21 individual case reports (Table 1). No controlled trials were found. Individuals reported in the studies included 90 with partial lipodystrophy (72 due to *LMNA* mutation and 15 due to *PPARG* mutation), 42 with generalized lipodystrophy (21 *AGPAT2*, 21 *BSCL2*, 2 *LMNA*), and 17 with IR due to *INSR* mutation(s). Among the interventions described, only the responses to metreleptin (111 recipients), thiazolidinediones (13 recipients) and rhIGF-1 (alone or as a composite with IGFBP3) (15 recipients) were described in more than 5 cases (Table 1). This meant that for the large preponderance of possible genotype-treatment combinations no specific data were recovered (Supplemental Table 3). Full outcome data extracted are summarised in Supplemental Table 4, and subject-level data are shown in Supplemental Figures 1 through 8.

**Table 1:** Summary characteristics of included studies. *Based on NHLBI quality assessment tool; ^#^Numbers in brackets are for partial lipodystrophy/generalised lipodystrophy/ insulin receptor individuals respectively. Abbreviations: rhIGF-1, recombinant human insulin-like growth factor 1; IGFBP3, insulin-like growth factor binding protein 3; SGLT2i, sodium-glucose co-transporter-2 inhibitor

| **Study types** | **Number of studies** |
| --- | --- |
| Case reports | 21 |
| Non-randomised experimental study | 10 |
| Case series | 8 |
| **Study Quality\*** | **Number of studies** |
| Good | 0 |
| Fair | 15 |
| Poor | 28 |
| **Phenotypes** | **Number of participants** |
| Partial lipodystrophy | 90<br>(72 *LMNA*, 15 *PPARG*, 2 *PLIN1*, 1 *PIK3R1*) |
| Generalised lipodystrophy | 56<br>(21 *AGPAT2*, 21 *BSCL2*, 1 *PTRF*, 2 *LMNA*) |
| Insulin receptor | 17 (7 Monoallelic, 10 Biallelic) |
| **Intervention** | **Number of participants** |
| Metreleptin | 111 (71/40/0) |
| rhIGF-1 or rhIGF-1/IGFBP3 composite | 15 (0/0/15) |
| Thiazolidinedione | 13 (12/1/0) |
| Metformin | 5 (2/1/2) |
| Bariatric surgery | 4 (4/0/0) |
| SGLT2i | 2 (1/1/0) |

### Metreleptin treatment was associated with improved metabolic control in lipodystrophy

In our registered systematic review plan we posed several subquestions about treatment of monogenic IR subtypes that we felt were tractable. The first related to the risks and benefits (assessed by side effects, A1c, serum triglyceride concentration, body mass index (BMI), and indices of fatty liver) of metreleptin in patients with different monogenic subtypes of lipodystrophy. The response to metreleptin was described in 111 people (71 with partial lipodystrophy, 40 with generalized lipodystrophy) ^12–22^. Metreleptin was administered for 19±20 months (median 12, range 1-108) and was associated with lowering of A1c in aggregated lipodystrophy, in generalized and partial subgroups, and in all genetic subgroups for whom sufficient patients were reported, namely those with *LMNA*, *PPARG*, *AGPAT2* and *BSCL2* mutations (0.5 to 1.5% least square mean reduction) (Level 3 evidence, Supplemental Table 4, Figure 2). Metreleptin treatment was also associated with lowering of serum triglyceride concentration in aggregated lipodystrophy, in generalized and partial subgroups, and in those with *LMNA*, *PPARG*, *AGPAT2* and *BSCL2* mutations (92 to 1760 mg/dL least square mean reduction for analyses of untransformed data) (Level 3 evidence, Supplemental Table 4, Figure 2). BMI was lower after treatment in aggregated lipodystrophy, in generalized and partial subgroups, and in those with *LMNA or BSCL2* mutations, but not *PPARG* or *AGPAT2* mutations (Level 3 evidence, Supplemental Table 4, Figure 2). Liver outcomes reported were too heterogeneous to analyse in aggregate. Only a single adverse event, namely hypoglycemia, was reported.

**Figure 2:**
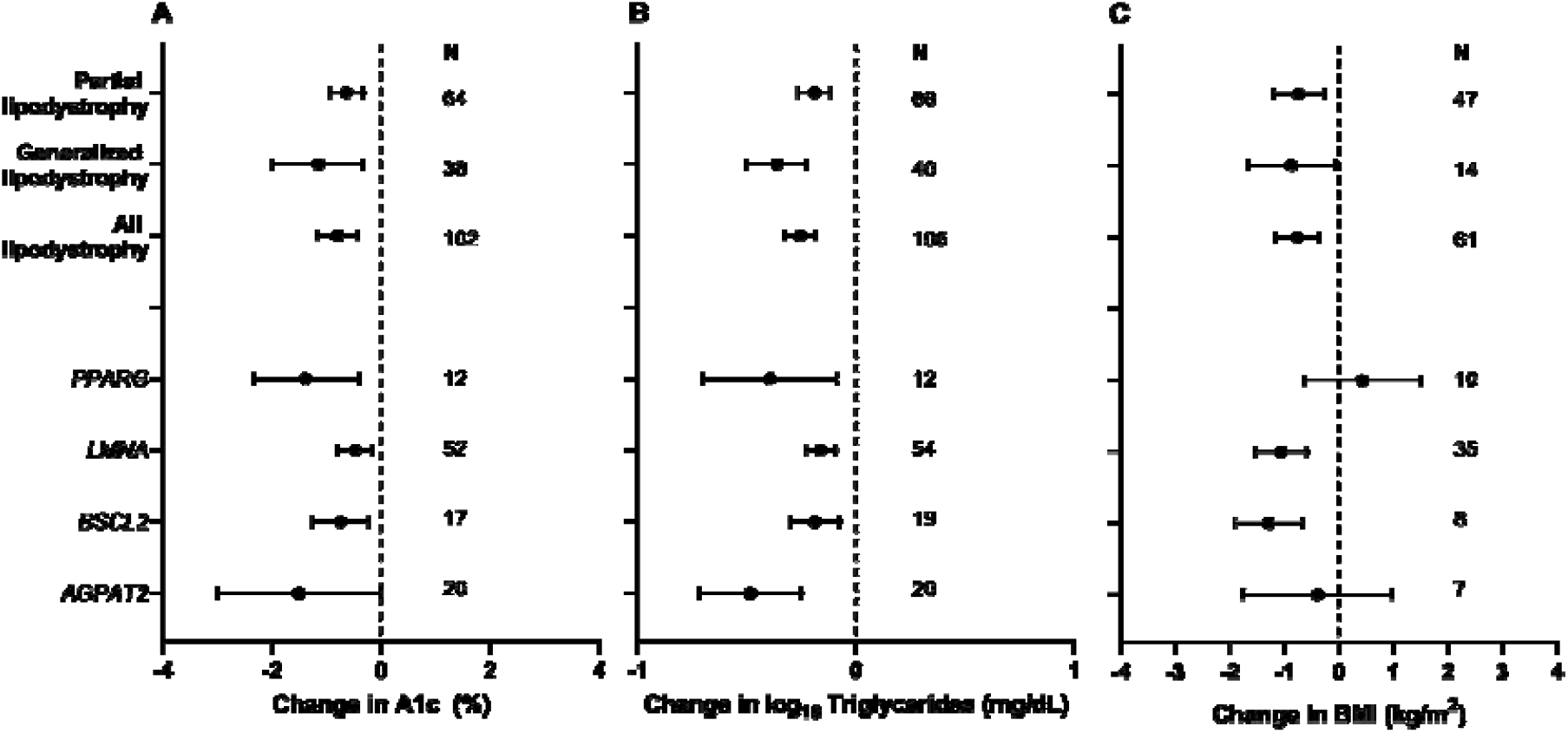
Effects of metreleptin in monogenic forms of lipodystrophy. Least square mean change in (A) Hemoglobin A1c (A1c), (B) Log_10_ serum triglyceride concentration and (C) Body Mass Index (BMI) in patients with partial lipodystrophy, generalized lipodystrophy, all forms of lipodystrophy, and subgroups with *PPARG*, *LMNA*, *BSCL2*, and *AGPAT2* mutations.

### Thiazolidinedione treatment showed variable efficacy in limited studies

We next addressed the evidence of risks and benefits of thizolidinediones (TZDs) in patients with lipodystrophy. We were specifically interested in any evidence of a greater or lesser response in partial lipodystrophy caused by *PPARG* variants than in other lipodystrophy subtypes, as TZDs are potent ligands for the gene product of the *PPARG* gene, the master regulator of adipocyte differentiation. The response to TZDs was described in only 13 people, however (12 FPLD, 1 CGL) ^23–32^. TZDs were administered for 29±28 months (median 24, range 2-96). TZD use was associated with improved A1c in aggregated lipodystrophy (least square mean reduction 2.2%) and in *PPARG*-related but not *LMNA*-related partial lipodystrophy (Level 4 evidence, Supplemental Table 4, Figure 3). Serum triglyceride concentration decreased in aggregated lipodystrophy and in those with *LMNA*-related but not *PPARG*-related partial lipodystrophy (Level 4 evidence, Supplemental Table 4, Figure 3). No adverse events were reported.

**Figure 3:**
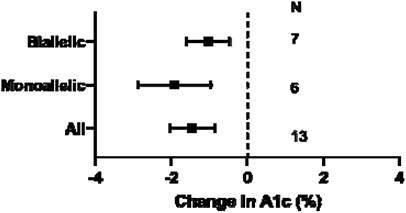
Effects of recombinant human Insulin-like Growth Factor-1 (rhIGF) alone or in combination with Insulin-like Growth Factor Binding Protein-3 (IGFBP3) in patients with *INSR* mutations. Least square mean change in hemoglobin A1c (A1c), in all patients with *INSR* mutations, and in subgroups with biallelic and monoallelic mutations.

### rhIGF-1 treatment in *INSR*-related IR was associated with improvement in A1c

Our last specific question related to the risks (e.g. tumors, hypoglycemia, cardiac hypertrophy, other soft tissue overgrowth) and benefits (assessed by A1c) of recombinant human IGF-1 (rhIGF-1) or IGF-1/IGFBP3 composite in patients with pathogenic *INSR* variants. The response to rhIGF-1 was described in 15 people with pathogenic *INSR* variants for a mean of 50±86 months (median 12, range 1-288) ^33–42^. In *INSR*-related IR, we found that use of rhIGF-1, alone or as a composite with IGFBP3, was associated with improvement in A1c, and this was true also in subgroups with monoallelic and biallelic variants (1 to 2% least square mean reduction, Level 4 evidence, Supplemental Table 4, Figure 4). One instance of increased soft tissue overgrowth and two episodes of hypoglycemia was reported.

**Figure 4:**
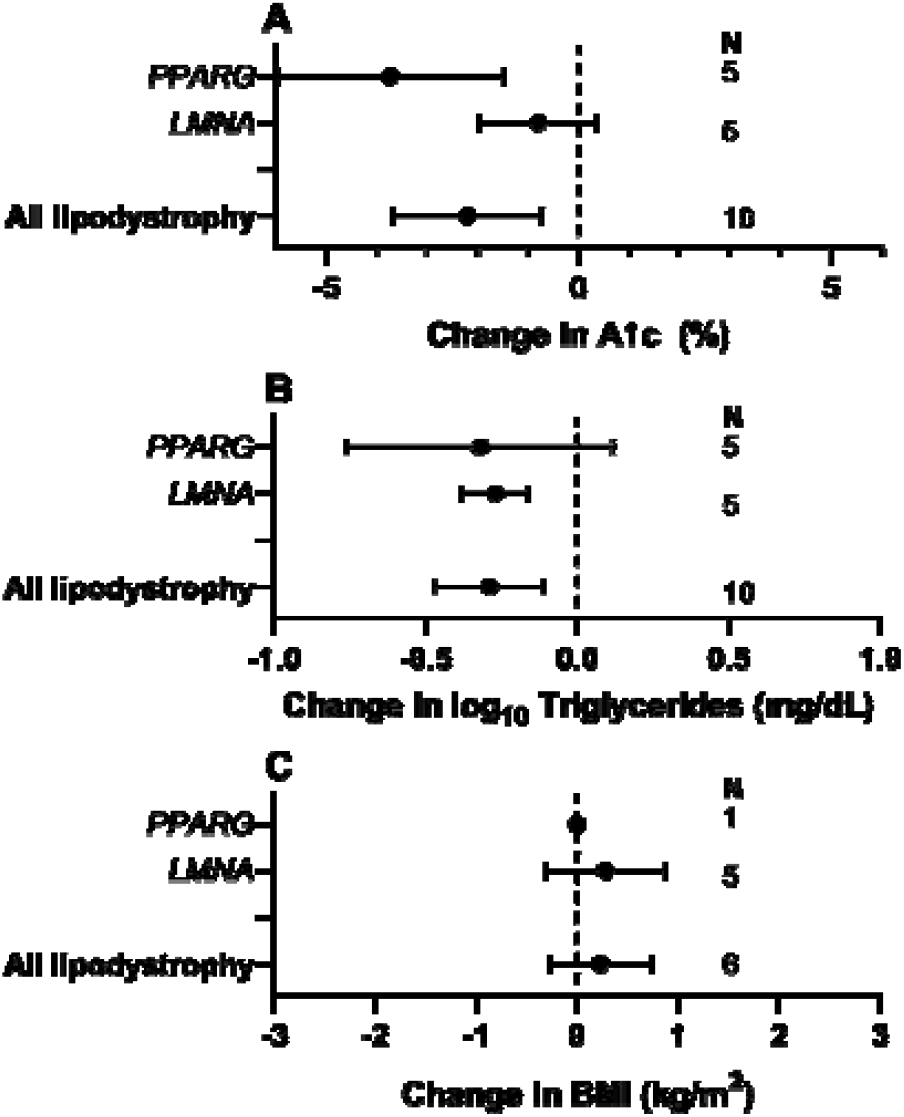
Effects of thiazolidinediones in monogenic forms of lipodystrophy. Least square mean change in (A) Hemoglobin A1c (A1c), (B) Log_10_ serum triglyceride concentration and (C) Body Mass Index (BMI) in patients with partial lipodystrophy, generalized lipodystrophy, all forms of lipodystrophy, and subgroups with *PPARG*, and *LMNA* mutations.

### Many questions about genotype-stratified treatment were not addressed

While many other interesting and clinically relevant questions arise about other potential genotype-specific responses to therapy in monogenic IR, the small size or absence of other genotype by treatment groups precluded the drawing of conclusions about risks and benefits, including for very widely used medications such as metformin ^25, 43–45^, newer agents commonly used in type 2 diabetes including SGLT2 inhibitors ^46, 47^ and GLP1 agonists, and non pharmacologic interventions such as bariatric surgery ^48–50^.

## Discussion

Thirty-five years since *INSR* mutations were identified in extreme IR ^51, 52^, and 23 years since the first monogenic cause of lipodystrophy was reported 53, many different forms of monogenic IR are known ^1–3, 54^. These are associated with substantial early morbidity and mortality, ranging from death in infancy to accelerated complications of diabetes and fatty liver disease in adulthood, depending on the genetic subtype. Several opportunities for genotype-guided, targeted treatment are suggested by the causal genes, and so we set out to review the current evidence guiding treatment of monogenic IR stratified by genetic aetiology. We found a paucity of high-quality evidence (all level 3 to 4). No controlled trials of any intervention were identified, and there was substantial heterogeneity of study populations and intervention regimens, even for the same interventional agent.

The evidence which we did find, from a small number of uncontrolled experimental studies, augmented by case series and numerous case reports, suggest that metreleptin offers metabolic benefits across different lipodystrophy subtypes, in keeping with its licensing for use in some patients with lipodystrophy in both Europe and the USA. Notably, the evidence base considered by licensing authorities was larger than the one we present, including many studies of phenotypically ascertained lipodystrophy that included acquired or idiopathic disease. In contrast we have addressed solely individuals with lipodystrophy caused by variation in a single gene. The limited data we identified do not clearly support differential effects among different monogenic lipodystrophy subgroups, but for many subtypes numbers reported are very small. Moreover although responses appear comparable for partial and generalised lipodystrophy, this is highly likely to reflect selection bias in studies of partial lipodystrophy towards those with more severe metabolic complications and lower baseline serum leptin concentrations.

A clear opportunity for precision diabetes therapy in monogenic IR is offered by the IR and lipodystrophy caused by mutations in *PPARG*, which encodes the target for thiazolidinediones (TZDs) such as pioglitazone ^55, 56^. PPARG is a nuclear receptor that serves as the master transcriptional driver of adipocyte differentiation, and so as soon as *PPARG* mutations were identified to cause severe IR, there was interest in the potential of TZDs as specific treatments. Although we found small scale evidence supporting greater A1c reduction with TZDs in *PPARG* vs *LMNA*-related lipodystrophy, only 5 patients with *PPARG*-related lipodystrophy in whom TZD effects were clearly described were reported, and responses were inconsistent. Thus, it remains unclear whether people with IR due to *PPARG* variants are more or indeed less sensitive to TZDs than people with other forms of lipodystrophy. Loss-of-function *PPARG* mutations are the second commonest cause of familial partial lipodystrophy 2, and the function of coding missense variants in *PPARG* has been assayed systematically to accelerate genetic diagnosis 57, so the opportunity to test genotype-related therapy in *PPARG*-related IR seems particularly tractable in future.

Other obvious questions about targeted treatment of monogenic, lipodystrophic IR are not addressed by current evidence. Important examples relate to the risks and benefits of treatments used in type 2 diabetes such as GLP-1 agonists and SGLT2 inhibitors. It is rational to suppose that these medications, which decrease weight as well as improving glycaemia in those with raised BMI and diabetes, may also be efficacious in lipodystrophy even where BMI is normal or only slightly raised. This is because in both situations adipose storage capacity is exceeded, leading to “fat failure”. It is the offloading of overloaded adipose tissue, rather than the baseline BMI/adipose mass, which underlies the efficacy of therapy. However GLP-1 agonists are contraindicated in those with prior pancreatitis, while SGLT2 inhibitor use can be complicated by diabetic ketoacidosis. In untreated lipodystrophy pancreatitis is common, yet this is due to hypertriglyceridaemia, which is likely to be improved by GLP-1 agonist use, while excessive supply of free fatty acids to the liver may promote ketogenesis. Thus assessment of both classes of drug in lipodystrophy and its genetic subgroups will be important to quantify risks and benefits, which may be distinct to those in obesity-related diabetes.

A further question we prespecified related to the use of rhIGF1 in people with severe IR due to *INSR* mutations. This use of rhIGF-1 was first described in recessive *INSR* defects in the early 1990s 42, based on the rationale that IGF-1 activates a receptor and signalling pathway very closely similar to those activated by insulin. Based on case reports, case series and narrative reviews, rhIGF-1 is now commonly used in neonates with extreme IR due to biallelic *INSR* mutations, although, unlike metreleptin in lipodystrophy, this use is still unlicensed. Our review of published data is consistent with glycaemic benefits of rhIGF-1, alone or in composite form with its binding protein IGFBP3, in *INSR* mutations. Nevertheless, such studies are challenging to interpret and are potentially fraught with bias of different types, particularly publication bias favouring positive outcomes. Responses to rhIGF1 are also challenging to determine in uncontrolled studies as small differences in residual function of mutated receptors can have substantial effects on the severity and natural history of the resulting IR, yet relatively few *INSR* mutations have been studied functionally. This underlines the narrow nature of, and significant residual uncertainty in, the evidence base for use of rhIGF-1 in monogenic IR.

There are several reasons why important questions about precision treatment of monogenic IR have not been settled. Although severe autosomal recessive IR is usually detected in infancy, commoner dominant forms of monogenic IR are often diagnosed relatively late, often only after years of management based on presumptive diagnoses of type 2 or sometimes type 1 diabetes. Initial management as type 2 diabetes means that by the time a clinical and then genetic diagnosis is made, most patents have been treated with agents such as metformin, and increasingly SGLT2 inhibitors or GLP-1 agonists, outside trial settings. It is not clear that harm is caused by such use of drugs with well-established safety profiles and efficacy in type 2 diabetes, but the lack of systematic data gathering precludes identification of specific drug-genotype interactions. Moreover, because attempts to gather evidence for monogenic IR treatment has tended to focus on high-cost adjunctive therapies such as metreleptin, the evidence base for their use is better developed, although controlled trials are lacking. Licensing of high-cost treatments such as metreleptin in lipodystrophy, while effects of many more commonly used, cheaper drugs with well-established safety profiles lack formal testing in monogenic IR is potentially problematic, skewing incentives and guidelines towards expensive therapy before optimal treatment algorithms have been established.

Other challenges in conducting trials in monogenic IR arise from the exquisite sensitivity of IR to exacerbating factors such as puberty, diet, and energy balance. This creates a “signal to noise” problem particularly problematic in uncontrolled studies, in which non-pharmacological components of interventions such as increased support for behavioural change may confound attribution of beneficial outcomes to pharmacological agents tested.

The key question now is how the evidence base for managing monogenic severe IR can be improved in the face of constraints in studying rare, clinically heterogeneous, and geographically dispersed patients who are often diagnosed late with a condition that is exquisitely environmentally sensitive. Growing interest in and development of methodologies for clinical trials in rare disease 58, including Bayesian methodologies ^59, 60^, and hybrid single- and multi-site designs 61 offer hope for future filling of evidence gaps. One important and pragmatic opportunity arises from the development of large regional, national and international networks and registries for lipodystrophy (e.g. the Europe-based ECLip registry 62), allied to emergence of randomised registry-based trial (RRT) methodology ^63, 64^. RRTs have attracted increasing interest in several disease areas and are particularly suitable for evaluation of agents with well-established safety profiles. When a simple randomisation tool is deployed in the context of a registry, RRTs can offer rapid, cost-effective recruitment and high external validity (i.e. relevance to “real world” practice). In monogenic IR this would permit questions to be addressed about optimal usage of different “common” medications in different genetic subgroups, including the order of introduction of therapies, and their optimal combinations. The quality of such studies will critically rely on good registry design and quality and completeness of data capture ^63, 64^.

In summary, severe monogenic IR syndromes are clinically and genetically heterogeneous, with high early morbidity and mortality. However despite opportunities for targeted therapy of some monogenic subgroups based on the nature of the causal gene alteration, the evidence for genotype-stratified therapy is weak. This is in part because of the rarity and frequent late diagnosis of monogenic IR, but also because therapeutic research to date has focused largely on phenotypically ascertained cross cutting diagnoses such as lipodystrophy. We suggest that approaches such as RRTs hold the best hope to answer some of the persisting major questions about precision treatment in monogenic IR.

## Authors’ declaration of personal interests

R.K.S. has received speaker fees from Eli Lilly, Novo Nordisk, and Amryt. R. J. B. has received research support from Amryt, Third Rock Ventures, Ionis, and Regeneron. K.A.P. and S.A. report no conflicts of interest.

## Data Availability

Not applicable

## Acknowledgements

This research was funded in part, by the Wellcome Trust [Grant WT 210752 to RKS and WT 219606 to KAP]. For the purpose of open access, the author has applied a CC0 Public Domain Dedication to any Author Accepted Manuscript version arising from this submission. RJB and SA are supported by the intramural research program of the National Institute of Diabetes and Digestive and Kidney Diseases. The ADA/EASD Precision Diabetes Medicine Initiative, within which this work was conducted, has received the following support: The Covidence license was funded by Lund University (Sweden) for which technical support was provided by Maria Björklund and Krister Aronsson (Faculty of Medicine Library, Lund University, Sweden). Administrative support was provided by Lund University (Malmö, Sweden), University of Chicago (IL, USA), and the American Diabetes Association (Washington D.C., USA). The Novo Nordisk Foundation (Hellerup, Denmark) provided grant support for in-person writing group meetings (PI: L Phillipson, University of Chicago, IL).

## Supplemental Tables

**Supplemental Table 1:**
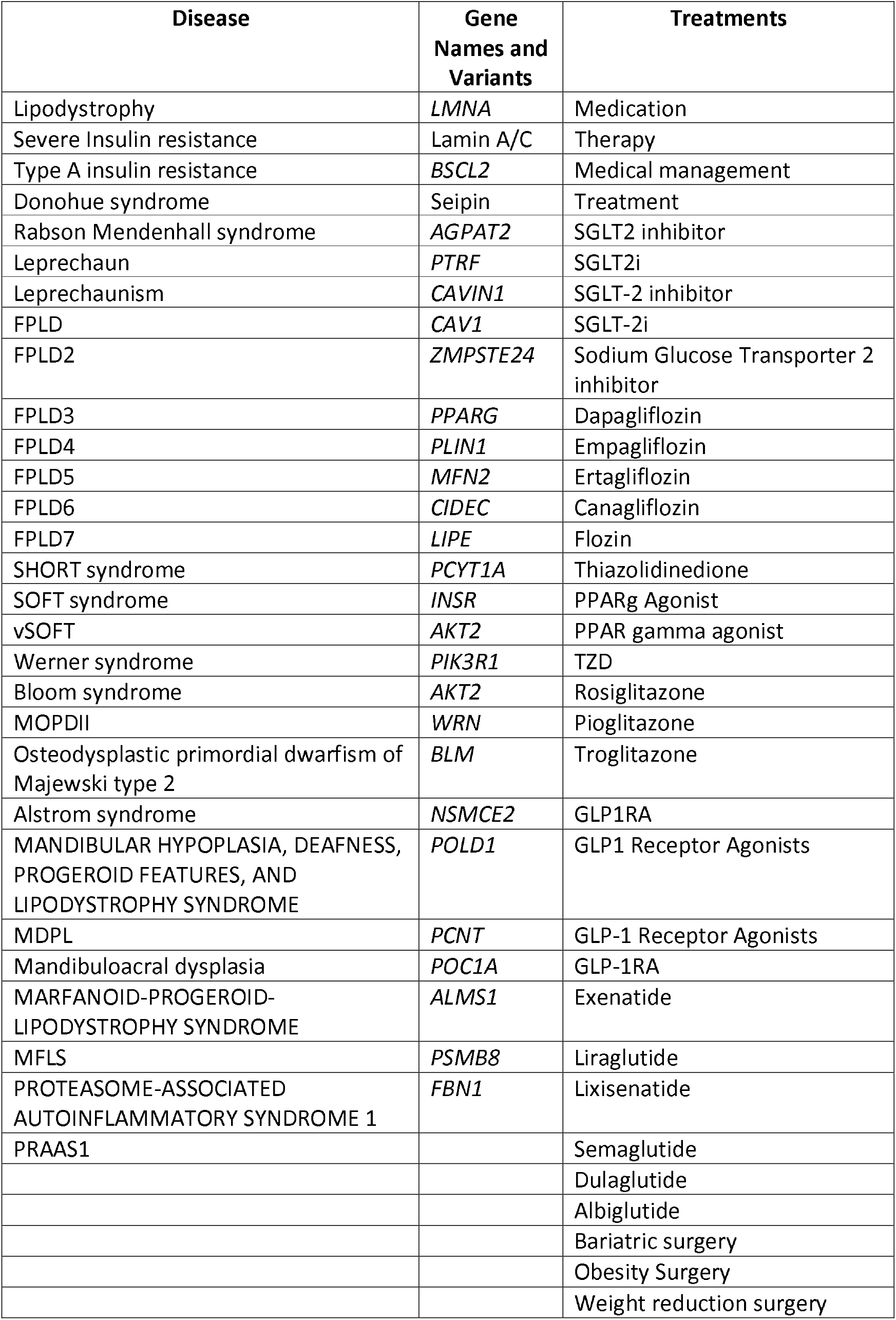
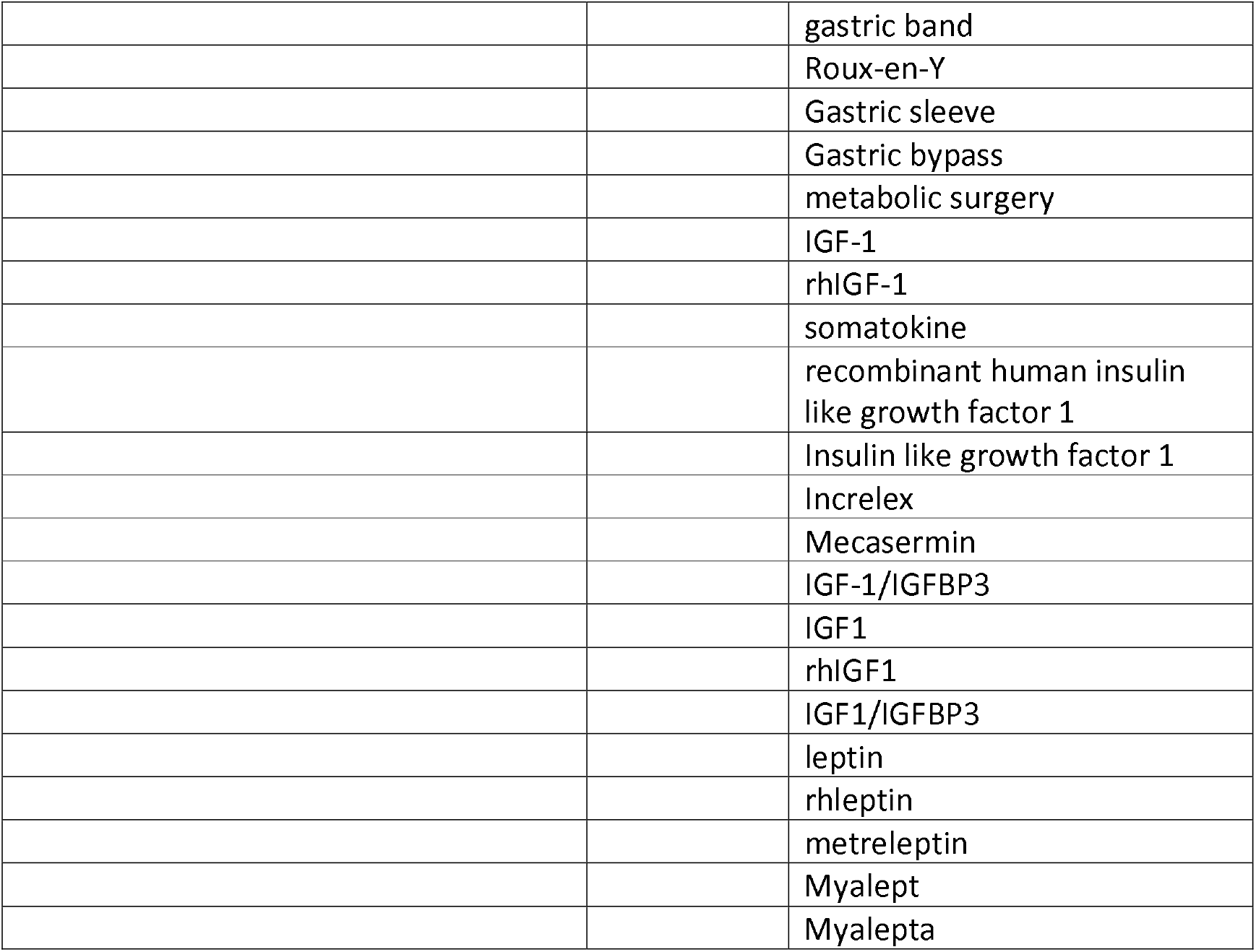
Descriptors of diseases, gene names and treatments included in the initial search.

**Supplemental Table 2:**
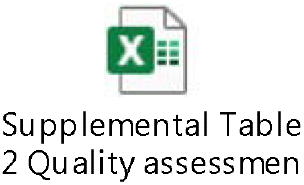
Detailed Quality Assessment of Included Studies.

**Supplemental Table 3:** Number of individuals by intervention and genotype in included studies. Abbreviations: rhIGF-1, recombinant human insulin-like growth factor 1; IGFBP3, insulin-like growth factor binding protein 3; SGLT2i, sodium-glucose co-transporter-2 inhibitor; TZD, thiazolidinedione; FPL = Familial Partial Lipodystrophy

|  |  | Treatment |  |  |  |  |  | Total |
| --- | --- | --- | --- | --- | --- | --- | --- | --- |
|  |  | rhIGF-1 or<br>rhIGF-1/IGFBP3<br>composite | SGLT2i | TZD | Bariatric<br>surgery | Metformin | Metreleptin |  |
| Gene | *AGPAT2* | 0 | 0 | 0 | 0 | 1 | 20 | 21 |
|  | *BSCL2* | 0 | 1 | 1 | 0 | 0 | 19 | 21 |
|  | *PTRF* | 0 | 0 | 0 | 0 | 0 | 1 | 1 |
|  | *LMNA*<br>(*progeroid*) | 0 | 0 | 0 | 0 | 0 | 2 | 2 |
|  | *LMNA*<br>(*FPL*) | 0 | 0 | 9 | 2 | 2 | 59 | 72 |
|  | *PPARG* | 0 | 0 | 5 | 0 | 0 | 10 | 15 |
|  | *PLIN1* | 0 | 0 | 0 | 2 | 0 | 0 | 2 |
|  | *PIK3R1* | 0 | 1 | 0 | 0 | 0 | 0 | 1 |
|  | *INSR* | 15 | 0 | 0 | 0 | 2 | 0 | 17 |
| *All partial LD* |  | -- | 1 | 14 | 4 | 2 | 71 | 90 |
| *All generalised LD* |  | -- | 1 | 1 | 0 | 1 | 40 | 43 |
| *All LD* |  | -- | 2 | 15 | 4 | 3 | 111 | 135 |

**Supplemental Table 4:**
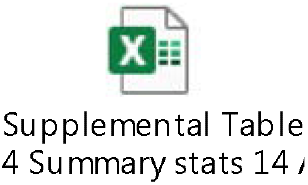
Summary statistics of extracted data.

## Supplemental Figures

**Supplemental Figure 1:**
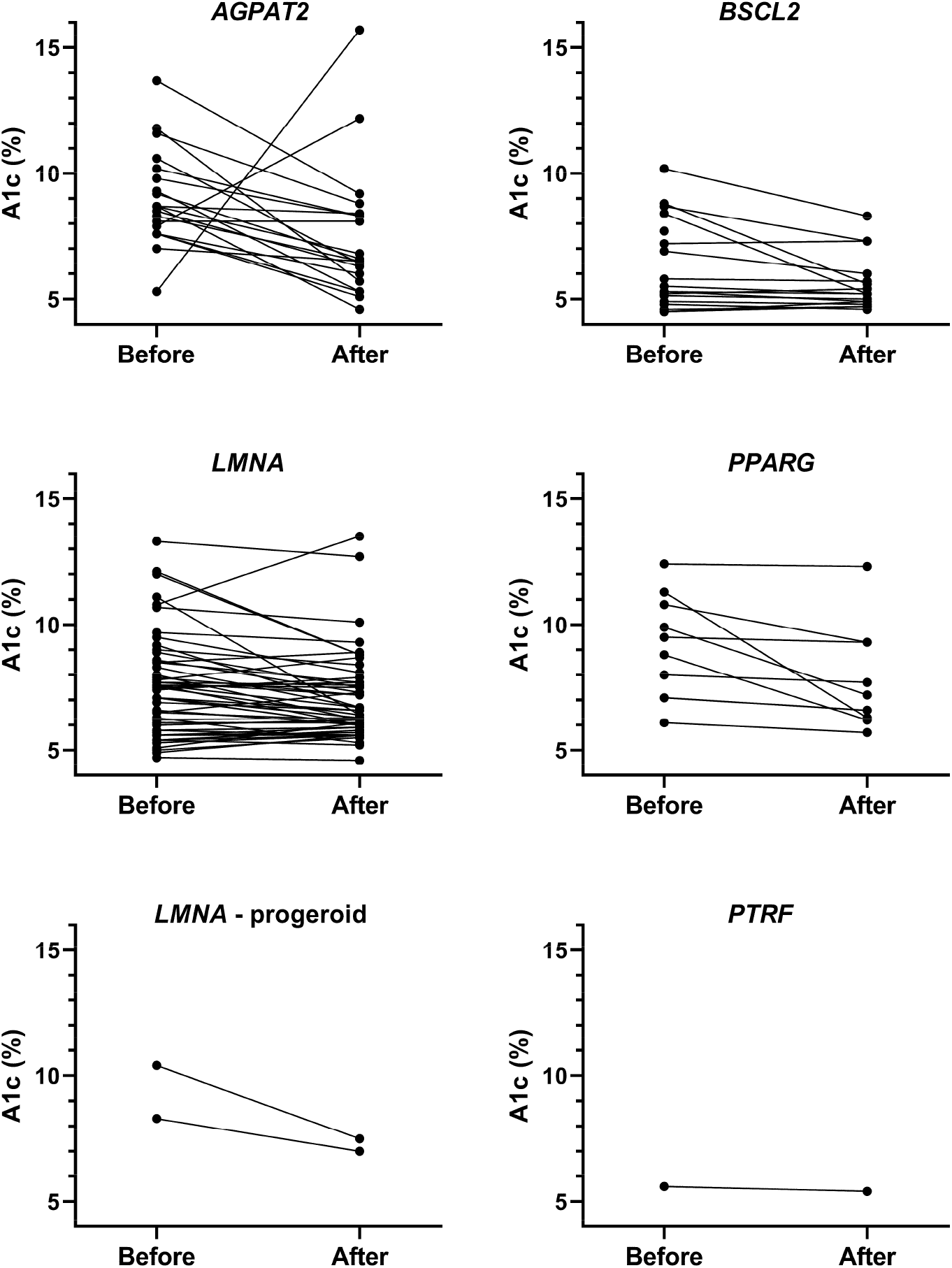
Effects of Metreleptin therapy on glycated haemoglobin (A1c) by genotype.

**Supplemental Figure 2:**
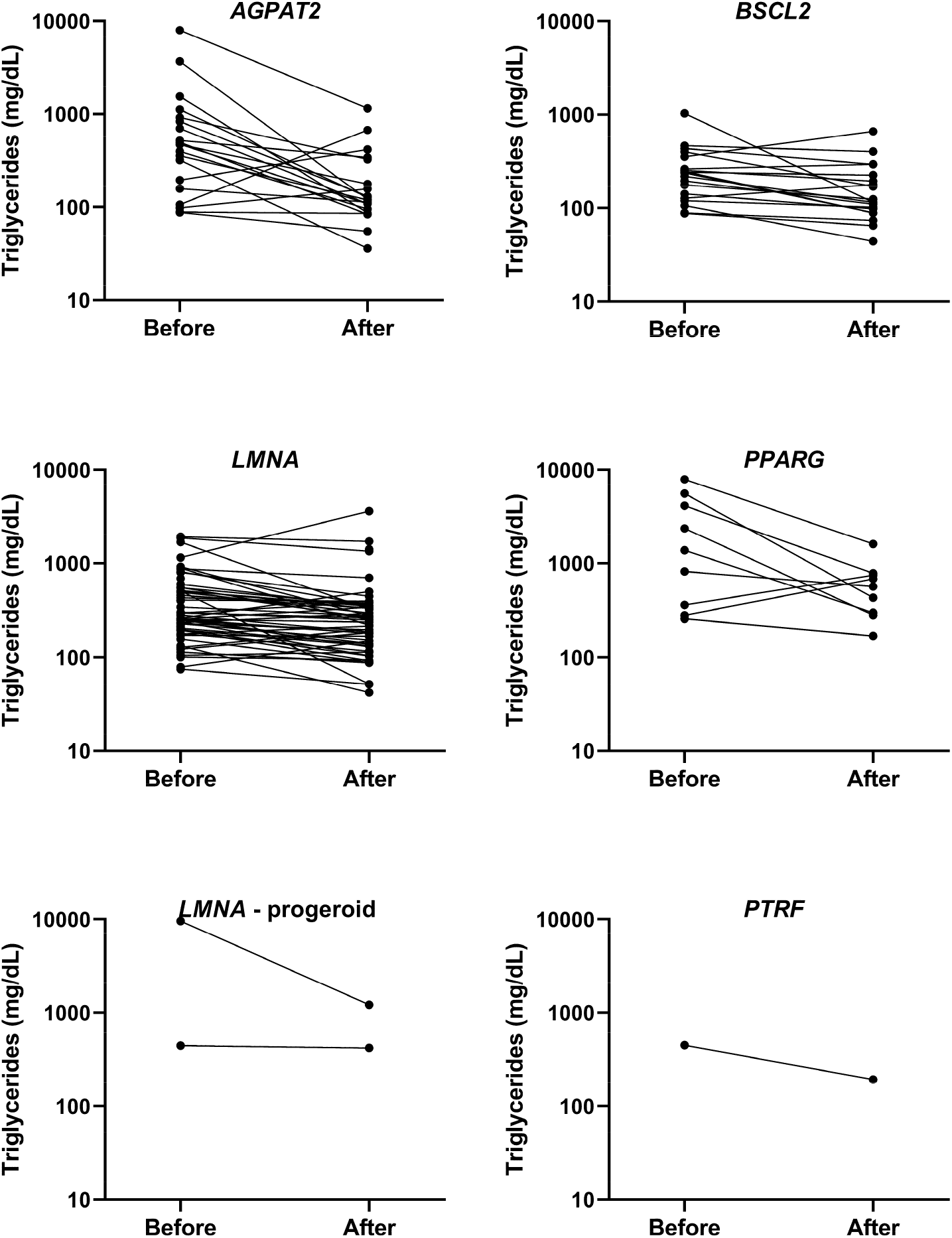
Effects of Metreleptin therapy on serum triglyceride concentration by genotype.

**Supplemental Figure 3:**
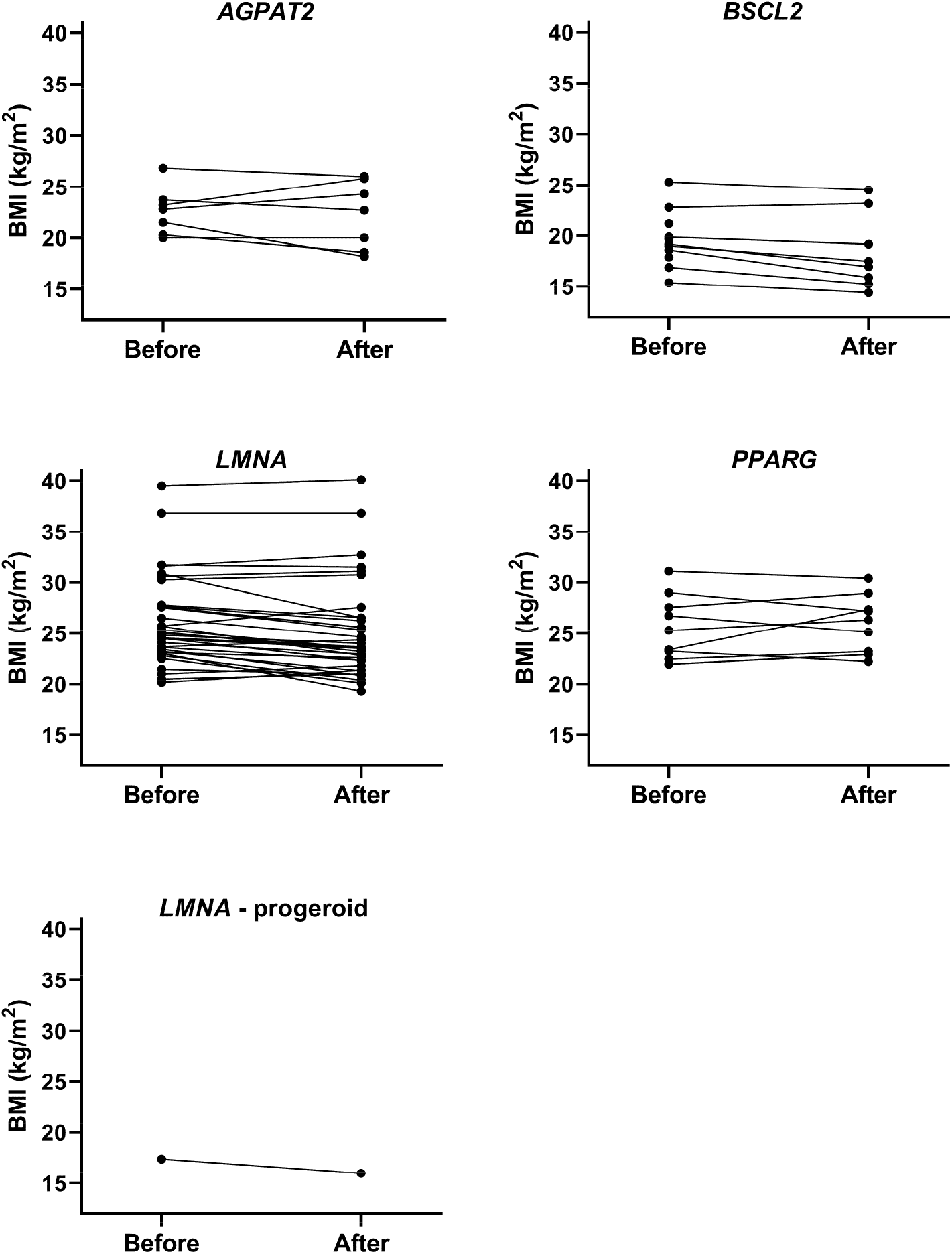
Effects of Metreleptin therapy on body mass index (BMI) by genotype.

**Supplemental Figure 4:**
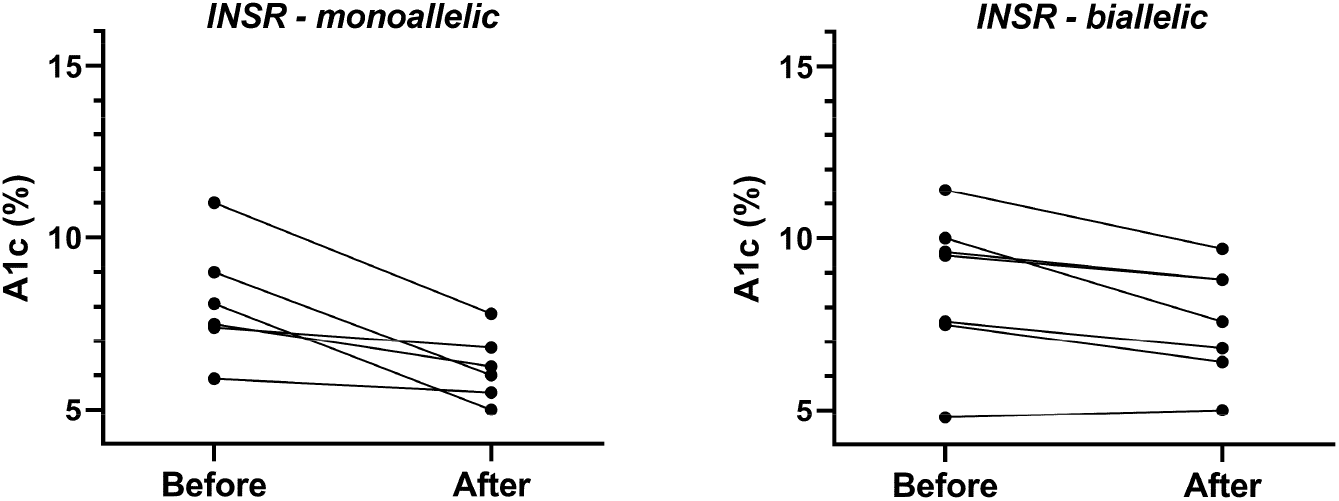
Effects of recombinant IGF-1 or IGF-1 plus IGFBP3 therapy on glycated haemoglobin (A1c) in monallelic and biallelic insulin receptoropathy.

**Supplemental Figure 5:**
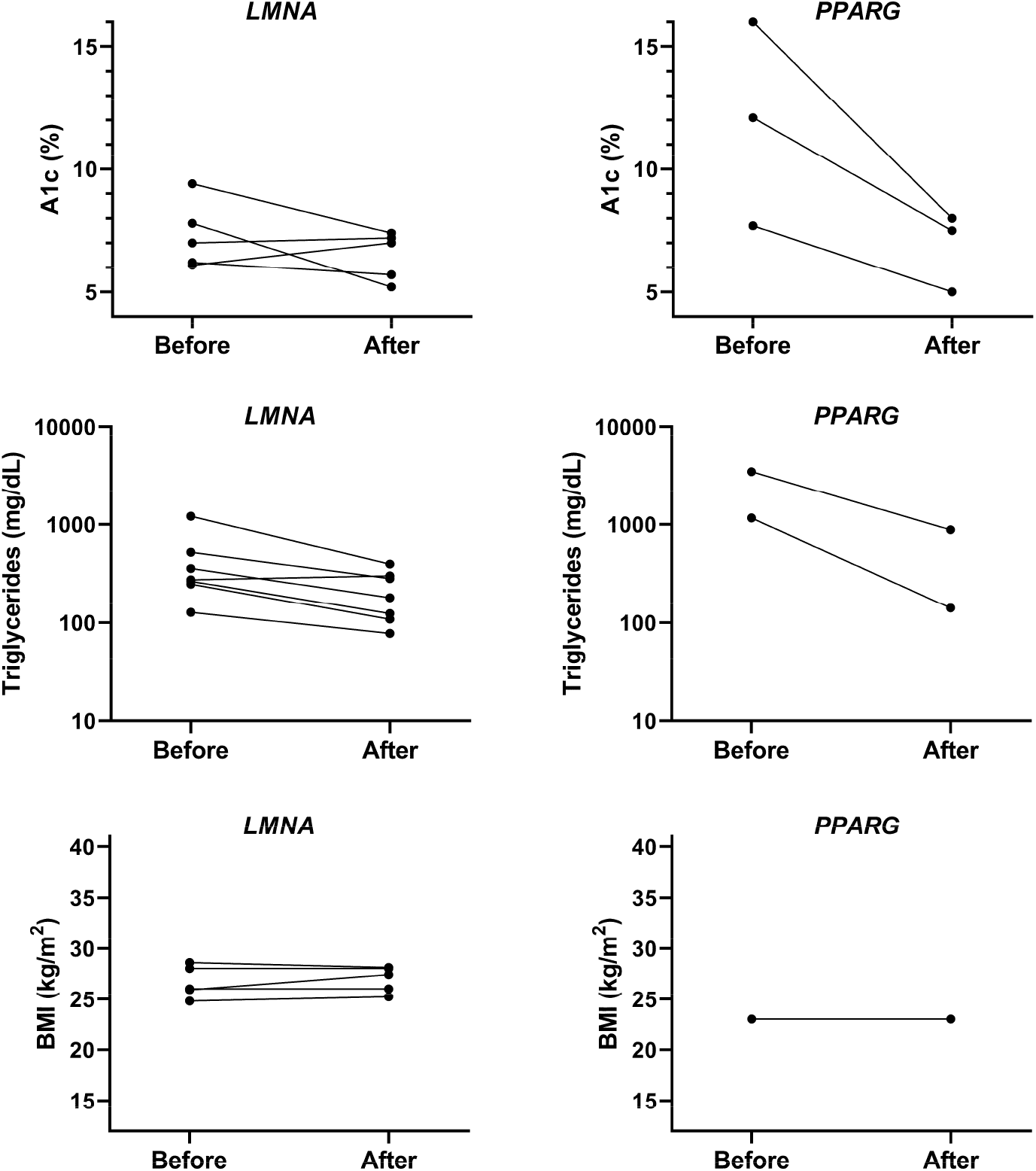
Effects of thiazolidinedione therapy by genotype.

**Supplemental Figure 6:**
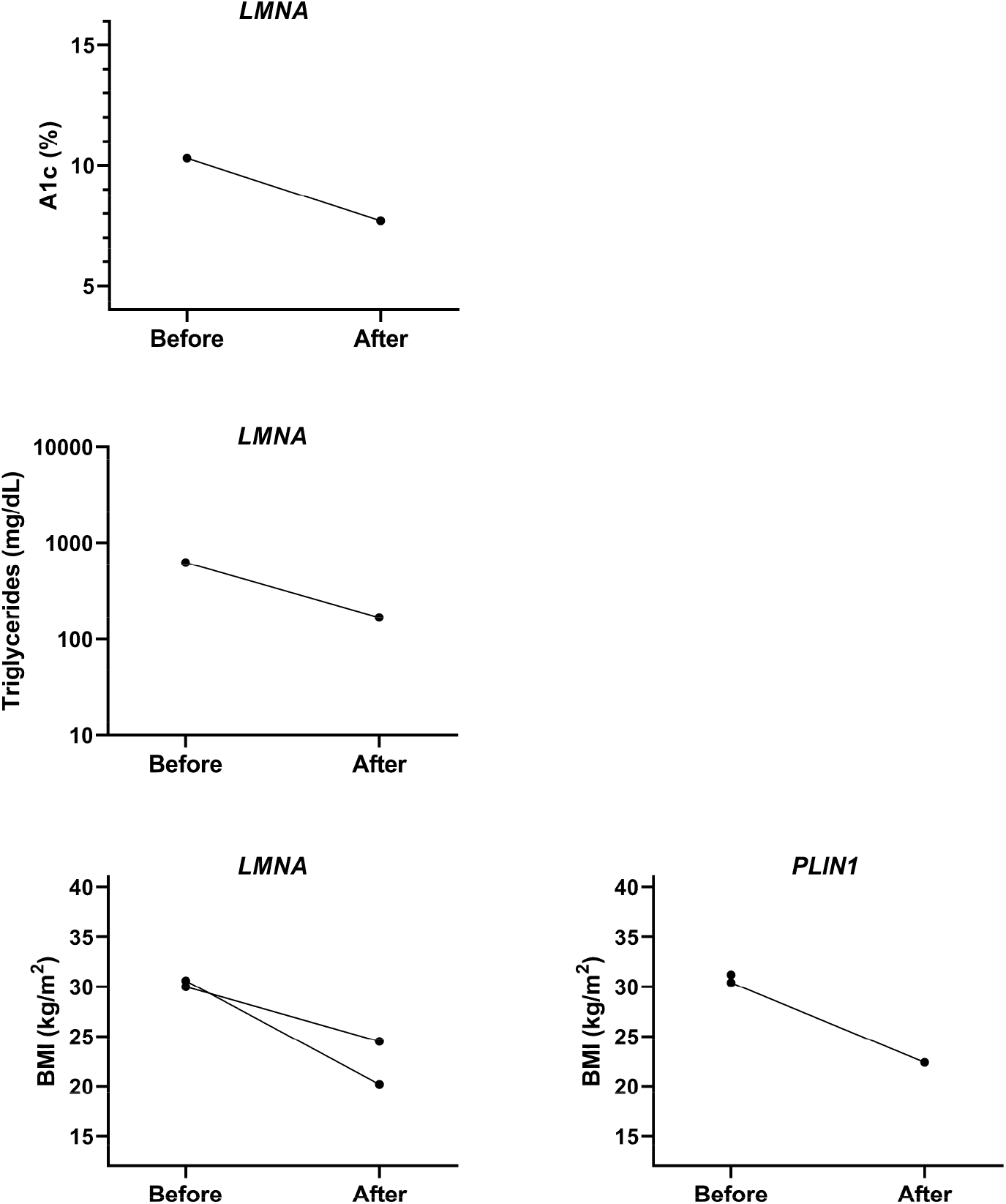
Effects of bariatric surgery by genotype.

**Supplemental Figure 7:**
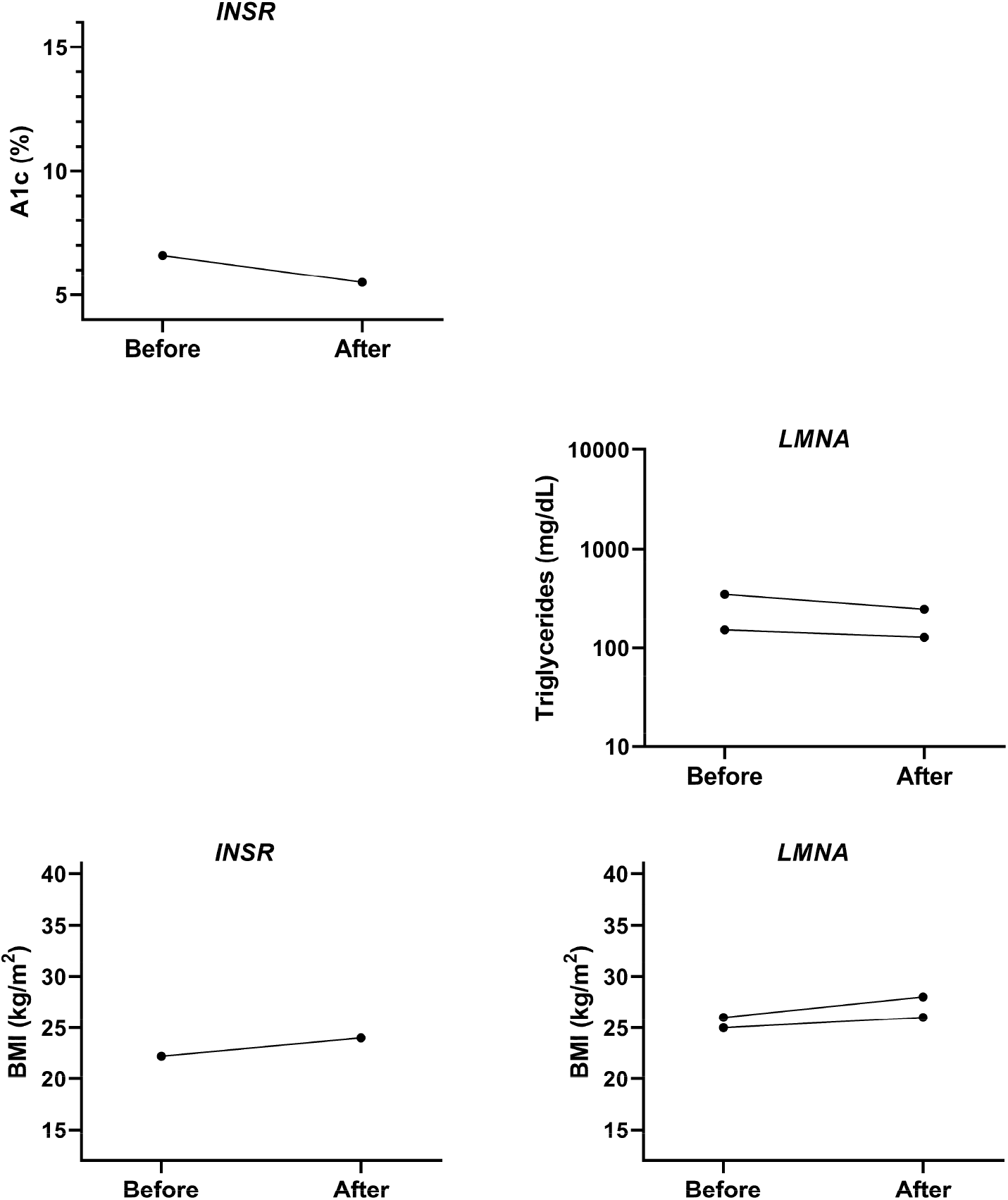
Effects of metformin therapy by genotype.

**Supplemental Figure 8:**
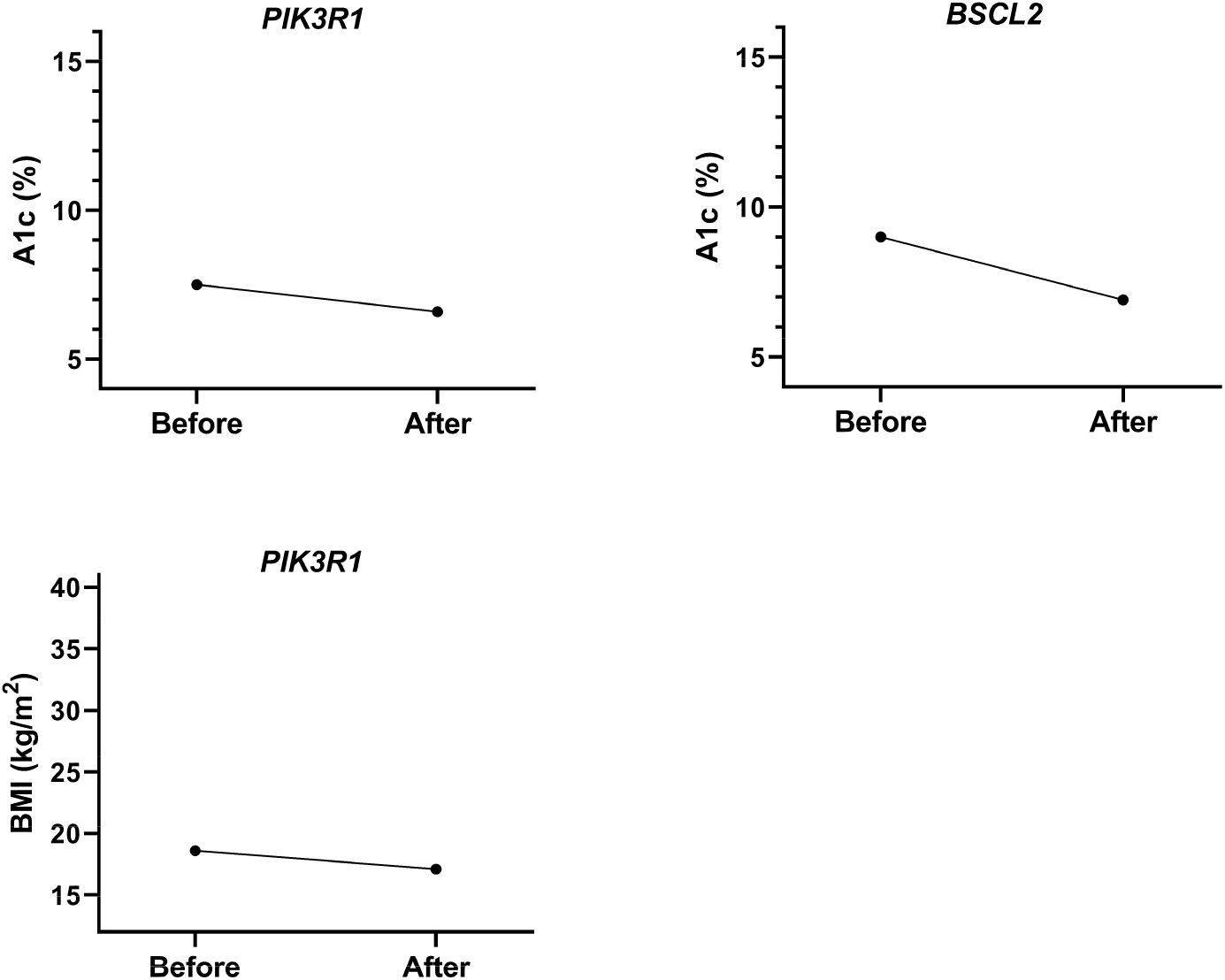
Effects of SGLT2 inhibitor therapy by genotype.

